# Real-Time fMRI Neurofeedback Targeting Cue Reactivity in Alcohol Use Disorder: Challenges and Insights from a Randomized Controlled Trial

**DOI:** 10.64898/2026.05.29.26354435

**Authors:** Patrick Halli, Franziska Weiss, Sarah Gerhardt, Jingying Zhang, Wolfgang H. Sommer, Falk Kiefer, Peter Kirsch, Martin Fungisai Gerchen

## Abstract

In a single-blind randomized controlled trial, we investigated the effectiveness of real-time fMRI neurofeedback delivered in 7 runs over three sessions across two weeks in N = 65 patients with alcohol use disorder. The intervention targeted modulation of ventral striatal cue reactivity to alcohol-related cues as well as enhancement of prefrontal control mechanisms in the right inferior frontal gyrus. The study design incorporate three experimental groups that either were instructed to downregulate a ventral striatum signal, upregulate the right inferior frontal gyrus, or upregulate negative functional connectivity between these two structures. In two active control groups participants were instructed to either up- or downregulate the primary auditory cortex.

We did not find an effect of ventral striatal downregulation or negative connectivity feedback, and a reduced striatal activation in the right inferior frontal gyrus upregulation group was accompanied by concurrent lower activation in the target structure, suggesting that our intended modulation approaches were not effective.

Identified problems that might have contributed to this unexpected outcome might have been the use of continuous feedback presentation that potentially confuses regulation target and reward processing in the ventral striatum, counterintuitive regulation directions, a lack of explicit strategy guidance and transparency about the targeted process, and generally the difficulty to recruit a sufficient number of eligible voluntary participants for a well-powered study with a complex design.

These insights emphasize the complex challenges of real-time fMRI neurofeedback interventions for the treatment of substance use disorders and could provide guidance for the development of more effective future approaches.

## Introduction

Alcohol is among the most lethal and socially burdensome substances, contributing substantially to disability-adjusted life years worldwide (MacKillop et al., 2022). While approximately 7% of all adult individuals worldwide develop an Alcohol Use Disorder (AUD), a significantly larger proportion experiences adverse consequences from alcohol consumption (World Health Organization 2024). Despite the availability of evidence-based treatment options - including psychotherapy, pharmacological interventions and social support services, a large number of individuals with AUD do not access formal treatment and those who do often experience considerable delays between disorder onset and treatment initiation. Moreover, AUD is a chronic, relapsing condition characterized by significant challenges in maintaining long-term abstinence (World Health Organization 2024), underscoring the need for novel treatment approaches. Although recent advances in basic neuroscience increasingly guide the design of interventions that target defined neural circuits, turning these insights into clinically effective treatments remains a challenge (Insel, 2012). One of the tools that has greatly advanced our understanding of the neural mechanisms underlying addiction is functional magnetic resonance imaging (fMRI). Building on this foundation, real-time fMRI neurofeedback (rtfMRI NF) leverages this technology to potentially modulate these neural processes in a therapeutic context (Dudek & Dodell-Feder, 2021; Pindi et al., 2022). In particular, rtfMRI NF aims to enable individuals to gain voluntary control over specific brain processes by providing real-time feedback of their own neural activity (Weiskopf, 2012).

Meta-analytic evidence suggests that rtfMRI NF can reliably induce target-specific modulation of brain activity. A comprehensive meta-analysis across psychiatric populations demonstrated robust regulation of trained regions both during active feedback and in transfer runs without feedback, while symptom-related improvements were typically small to moderate and showed substantial between-study variability (Dudek & Dodell-Feder, 2021). Similarly, another meta-analysis reported comparable clinical effects - particularly in affective disorders - but highlighted marked heterogeneity, predominantly small sample sizes, and methodological limitations across studies (Pindi et al., 2022). Complementary individual participant data analyses further indicate that successful self-regulation recruits a partly shared neural control network; however, stable predictors of learning success or clinical benefit remain elusive (Emmert et al., 2016; Haugg et al., 2020).

In the context of AUD rtfMRI NF has the potential to modulate neural structures associated with clinically relevant functions in AUD (Naqvi & Morgenstern, 2015), including ventral striatal cue reactivity and cognitive control, in individuals with AUD. The ventral striatum (VS), including the nucleus accumbens (NAcc), is a central hub of the limbic reward circuitry, mediating emotional reinforcement and playing a key role in the development and maintenance of addictive behaviors. Drugs of abuse prolong dopamine signaling in the NAcc and enhance dopaminergic activity in both the ventral tegmental area (VTA) and NAcc, thereby reshaping reward-related circuits (Koob & Volkow, 2016). In AUD, this circuitry can be triggered by alcohol-related cues - a phenomenon described as cue reactivity - which is associated with craving and relapse risk (Schacht et al., 2013). Recent meta-analytic evidence further suggests that craving, including cue-induced craving, reliably predicts future substance use and relapse, with these associations observed across both in vivo and pictorial cue exposure paradigms (Vafaie & Kober, 2022).

Regulating this reward-related hyperactivity requires the recruitment of executive control mechanisms. The right inferior frontal gyrus (rIFG), a core node of the executive control network, is consistently engaged during response inhibition and supports higher-order functions such as planning, volition and performance monitoring (Hampshire et al., 2010). Increased rIFG activation appears essential for effectively down-regulating VS activity, underscoring the functional interplay between these regions. In an fMRI NF study with heavy social drinkers, Kirsch et al. (2016) found that NF induced reduction of ventral striatal cue reactivity activation was accompanied by increased activation of the rIFG making this structure a promising NF target for reducing cue reactivity and enhancing inhibitory control.

Here, we conducted a randomized controlled trial to investigate the use of rtfMRI NF to downregulate ventral striatal cue reactivity in patients with AUD. By tailoring NF to clinically relevant circuits, the study aims to uncover effective strategies for restoring self-regulation, an essential capacity often compromised in AUD. We randomly assigned patients with AUD to one of three experimental or two control NF groups that, while seeing pictures of their preferred alcoholic beverages, either directly downregulated a VS feedback signal, upregulated a rIFG signal, or upregulated negative connectivity between these two structures. The two active control groups were instructed to upregulate or downregulate activity in the primary auditory cortex (Gerchen et al., 2018). We hypothesized that all three experimental groups will exhibit lower ventral striatal cue reactivity during NF in comparison to the respective control groups.

## Methods

### Sample

N = 65 patients diagnosed with AUD from both outpatient and inpatient clinics at the Department of Addiction Behavior and Addiction Medicine, Central Institute of Mental Health (CIMH), Mannheim, Germany were recruited. Participants were randomly assigned in a single-blind manner to one of five parallel groups, each receiving genuine rtfMRI NF targeting different brain processes (Figure 1). Monthly follow-up assessments were conducted over a period of three months.

**Figure 1.**
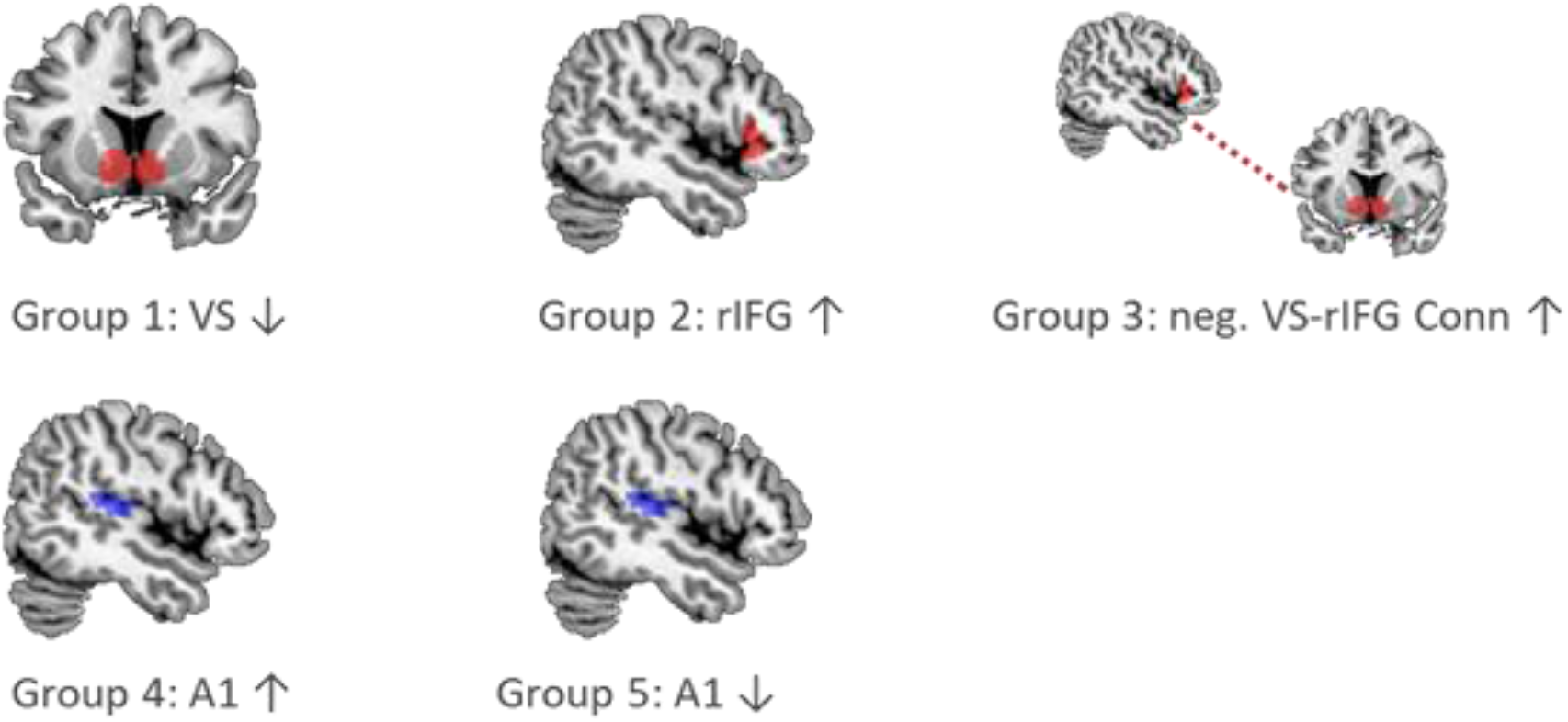
Experimental Groups. Participants were randomized into one of 5 experimental groups that all received rtfMRI NF. Group 1 received feedback from the VS and was instructed to downregulate the signal. Group 2 received feedback from the rIFG and was instructed to upregulate. Group 3 received a feedback signal representing the strength of negative functional connectivity between the VS and rIFG and was instructed to upregulate (i.e. increase negative connectivity). Group 4 and 5 were active control groups that up-(Group 4) or downregulated (Group 5) a signal from the right primary auditory cortex (A1). All participants were blind towards the feedback signal they received.

The study was registered in the German Clinical Trials Register (trial identifier: DRKS00010253; WHO Universal Trial Number (UTN): U1111–1181-4218) and conducted in compliance with the guidelines for Good Clinical Practice and the Declaration of Helsinki. The study protocol is described in Gerchen et al. (2018).

### Participant Eligibility and Recruitment

Participants were 18–65 years old, had normal or corrected-to-normal vision, met ICD-10 criteria for alcohol dependence (F10.2), and had maintained controlled abstinence for at least 5 days prior to inclusion. All had to have completed a medically supervised detoxification - including treatment of withdrawal symptoms with short-acting benzodiazepines or clomethiazole - which had to be finished at least three days before study enrollment. Exclusion criteria were any other Axis I psychiatric disorder (DSM-IV/ICD-10) in the past 12 months, except alcohol and nicotine use disorders and mild depressive symptoms related to alcohol consumption or detoxification, a positive urine drug screen, current use of psychotropic or anticonvulsant medication, epilepsy, neurological or severe medical illness, suicidal tendencies, pregnancy or breastfeeding. Recruitment was carried out by study team psychologists, who explained the study’s purpose and addressed any participant questions. Written informed consent was obtained before screening, and participants were informed that they could withdraw their consent at any time without providing a reason.

### MRI

#### NF Scanner

MRI scanning was conducted using a Siemens 3T Tim Trio scanner (Siemens Healthineers, Erlangen, Germany) at the CIMH in Mannheim, Germany. Each participant underwent NF training across three scanning sessions within a two-week period. On each scanning day, participants complete the following sequence: a 5:21-minute T1-weighted anatomical MPRAGE scan, a 12:00-minute resting-state functional scan acquired with closed eyes, and three NF runs, each lasting 9:29 minutes.

At the beginning of each session, the MPRAGE scan was acquired with the following parameters: repetition time (TR) = 2.3 s, echo time (TE) = 3.03 ms, flip angle = 9°, field of view = 256 mm, 192 sagittal slices, matrix size = 256 × 256, voxel size = 1 × 1 × 1 mm, using GRAPPA with iPAT = 2. Resting-state and NF runs were acquired using an echo-planar imaging (EPI) sequence with TR = 1.64 s, TE = 30 ms, flip angle = 73°, field of view = 192 mm, voxel size = 3 × 3 × 3 mm, 33 slices of 3 mm thickness, a distance factor of 33%, and GRAPPA with iPAT = 2. During functional imaging, respiration and pulse signals were acquired with built-in equipment.

#### NF Setup

The rtfMRI NF procedure is implemented using custom MATLAB scripts (MathWorks Inc., Sherborn, MA, USA) based on SPM functions (Wellcome Department of Cognitive Neurology, London, UK), in combination with Presentation software (Neurobehavioral Systems, Inc., Albany, CA, USA) for stimulus and feedback presentation. On each scan day, the anatomical image is first segmented and normalized to MNI space. The inverse deformation fields from normalization are then applied to warp region-of-interest (ROI) masks into native subject space.

During NF runs, reconstructed DICOM images were transferred from the scanner to a laptop for real-time preprocessing and feedback signal extraction using in-house software. Functional images were realigned to the mean of the first 10 volumes and resliced. ROI masks were resampled to match the current image space and mean intensity values from voxels within the target regions were extracted.

For activation-based NF, intensity values during the alcohol cue presentation were averaged over the last three volumes (moving average) to stabilize the signal and percent signal change was calculated relative to the preceding fixation block.

For connectivity-based NF, partial correlations between the VS and rIFG, adjusted for the cerebrospinal fluid signal, were computed over the preceding 15 volumes. In this exploratory condition, participants were instructed to increase the feedback signal, which represented the strength of negative connectivity between rIFG and VS in the upward direction. Importantly, a negative VS–rIFG correlation could also indicate increased VS activity during reduced prefrontal control. To ensure a top-down inhibitory configuration, the connectivity feedback algorithm includes additional constraints: connectivity-based feedback values were only displayed in the desired upregulation direction when VS activity decreased and rIFG activity increased; otherwise, values were inverted.

In both feedback types, the feedback scale was adaptively adjusted to reflect the maximum absolute signal change. The calculated feedback value was then transmitted to a presentation computer and displayed to the participant inside the scanner as a thermometer-style indicator next to an image of their preferred alcoholic beverage (beer, wine, or both). The thermometer updated with each new volume and the order of beverage images was randomized across participants.

#### NF Task

NF sessions followed a block design, alternating between the presentation of a fixation cross (baseline; 41 seconds) and an image of the participants preferred alcoholic beverage (beer, wine, or both; approximately 51 seconds). Each condition was presented six times per run. Approximately 10 seconds after the onset of the alcohol-related images, a feedback thermometer appeared on both sides of the picture and was updated at every TR (feedback duration: 41 seconds). On the first and third scanning session, the third run served as a transfer run without feedback, resulting in a total of seven NF runs.

On each scanning day, participants rated their alcohol craving using a visual analog scale immediately before and after the MRI session. Following the third NF session, participants completed a set of alcohol-related questionnaires.

### Statistical Analysis

To evaluate the effects of the rtfMRI NF, offline data analyses was performed using MATLAB and SPM 12. Preprocessing steps included slice-timing correction, realignment, anatomical segmentation, normalization to MNI space, spatial smoothing, and identification of outlier volumes (framewise displacement FD>0.5 mm, global intensity change z>4) due to excessive head motion.

The preprocessed data was subjected to first-level single-subject analyses, in which the time courses of the experimental conditions were convolved with the canonical hemodynamic response function and entered into a general linear model (GLM) to estimate brain activation during rtfMRI NF.

The GLM design matrix further contained the six standard motion parameters, dummy regressors marking motion outlier volumes and physiological nuisance regressors estimated from the recorded physiological signals with the PhysIO toolbox (Kasper et al., 2017). We conducted offline physiological signal-based correction of physiological artefacts, as described by Zhang et al. (2026).

First-level contrast images between NF and rest periods (NF > rest) were estimated and average activation was extracted from the NF target regions. These activation values were then compared between groups with two-sample t-tests implemented in a GLM with age and sex as covariates and the effect size Hedges’ g with its 90% confidence interval (CI) was estimated (see Gerchen et al., 2021). In these analyses, the experimental groups were compared with the respective up- or downregulation control group. Further, the combined experimental groups were compared with the combined control groups. Given the exploratory nature of the present analyses, we report results using uncorrected thresholds to maximize sensitivity to potential effects. Accordingly, 90% confidence intervals are presented, corresponding to a one-sided significance level of p < .05.

## Results

Our final sample included N=65 patients with AUD (see Table 1). In the main text, we report the results of the group comparisons, within-group effects on basic target process modulation can be found in the supplementary material (see Supplement Figures S1-S6 and Tables S13-S18).

**Table 1.** Descriptive Statistics for Demographic and Clinical Variables Across Neurofeedback Groups.

| Variable | VS down | rIFG up | rIFG–VS<br>connectivity<br>down | A1 up | A1 down | Overall | p |
| --- | --- | --- | --- | --- | --- | --- | --- |
| n | 13 | 14 | 12 | 9 | 17 | 65 | — |
| Age M (SD) | 43.15<br>(11.76) | 47.29<br>(8.63) | 49.17 (9.54) | 47.22 (6.24) | 47.35<br>(11.15) | 46.82 (9.83) | .641 |
| Sex (m/f) | 10 / 3 | 11 / 3 | 12 / 0 | 7 / 2 | 13 / 4 | 53 / 12 | .501 |
| BDI-Sum M (SD) | 11.42 (8.23) | 14.50<br>(12.07) | 10.70 (8.17) | 10.78 (8.73) | 14.00<br>(11.40) | 12.34 (9.64) | .839 |
| Baseline OCDS M (SD) | 23.08 (7.04) | 22.67<br>(7.72) | 25.18 (7.18) | 23.44 (4.07) | 24.50<br>(7.58) | 23.82 (6.84) | .901 |
| Drinking Days M (SD) | 70.31<br>(25.35) | 67.00<br>(15.59) | 66.67 (19.98) | 58.11<br>(23.85) | 67.69<br>(18.70) | 66.53<br>(20.28) | .736 |
| Cumulative Alcohol (g) M (SD) | 14315.69<br>(10990.23) | 14902.86<br>(7092.85) | 15168.00<br>(8468.48) | 13494.11<br>(19043.41) | 14642.06<br>(9359.89) | 14570.00<br>(10634.48) | .998 |

**Note.** Sex values represent male/female frequencies; BDI = Beck Depression Inventory; OCDS = Obsessive Compulsive Drinking Scale. Drinking days refer to the last 90 days; cumulative alcohol consumption refers to the total grams of alcohol consumed in the last 90 days. Continuous variables were compared using one-way analyses of variance (ANOVA); sex differences were tested using $\chi^2$ tests. Sample sizes vary across variables due to missing data (Age: 0; BDI: 12; OCDS: 5; drinking days: 1; cumulative alcohol consumption: 1).

### VS Modulation by NF

The central aim of our study is the reduction of ventral striatal cue reactivity, therefore we first investigated whether VS activation is downregulated by NF in the experimental groups in comparison to the respective control groups (Figure 2) over the 7 NF runs. When VS downregulation (group 1) was compared with A1 downregulation (group 5), no effect could be detected in any run. When VS activation during rIFG upregulation (group 2) and A1 upregulation (group 4) was compared, the rIFG upregulation group exhibited significant lower VS activation in the first 6 of the 7 NF runs at a nominally hypothesis-conform one-sided uncorrected significance threshold of p<0.05. The comparison between rIFG-VS negative connectivity upregulation (group 3) and A1 upregulation (group 4) showed nominally significant lower VS activation only during the first NF run. When all experimental groups (groups 1, 2 & 3) were compared to all control groups (groups 4 & 5), only in the first NF run nominally significant lower VS activation was found.

**Figure 2.**
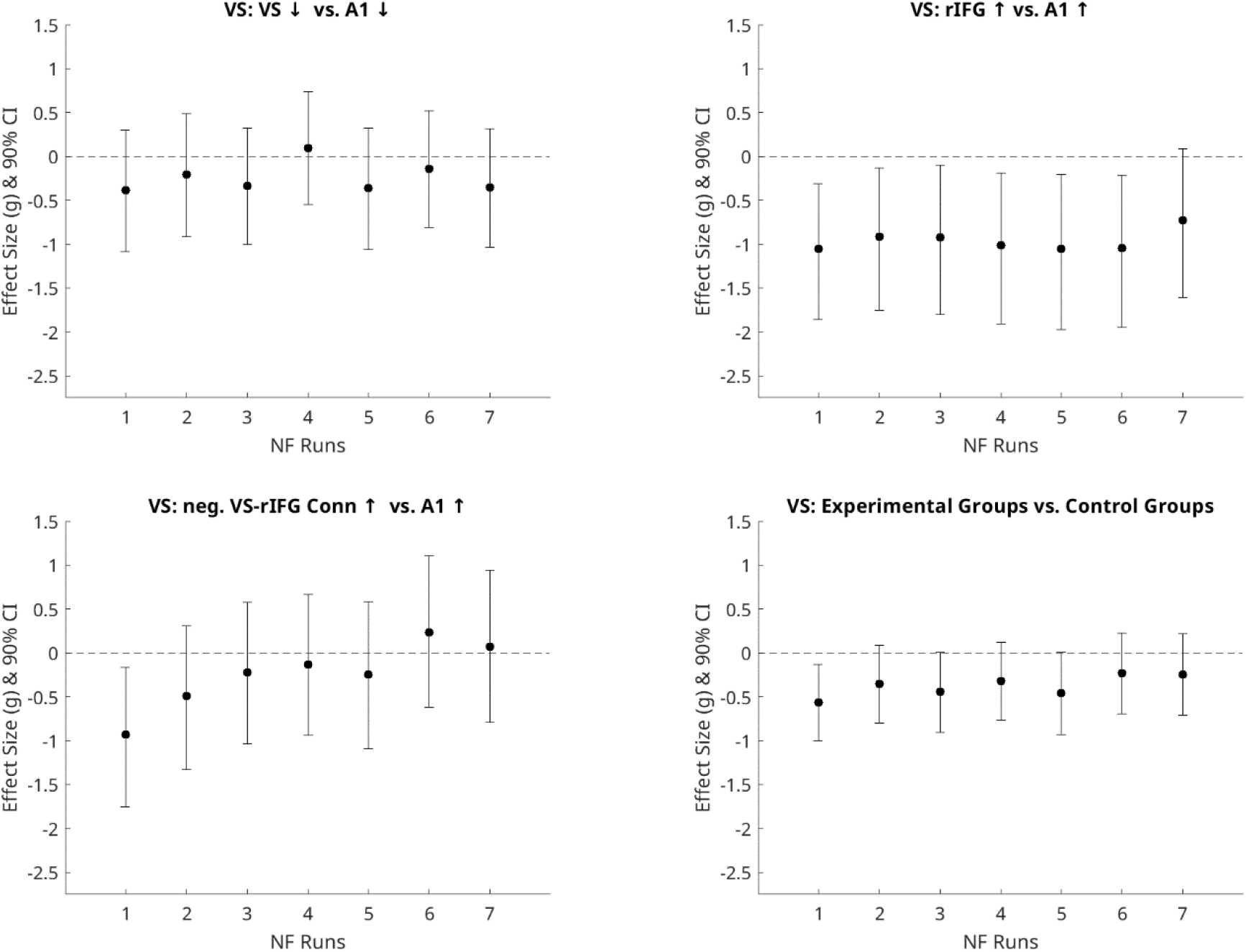
Group comparisons of NF effects in the VS. Effect sizes (Hedges’ g) with 90% confidence intervals (CI) for group comparisons of VS activation with two-sample t-tests across NF runs. Experimental groups are compared with the respective up- or downregulation control groups. In the lower right panel the combined comparison of the experimental groups (VS down / rIFG up / neg. VS-rIFG Conn up) versus the control groups (A1 up / A1 up) is shown. Negative values indicate a lower activation in the VS in the experimental groups during NF phases. Please note that the limits of the CI are equivalent to a one-sided uncorrected significance threshold of p<0.05. Exact test statistics for all analyses are provided in the Supplementary Material (Tables S1– S4)

### Changes in rIFG Activation during NF

Because we assumed that the rIFG plays a role in VS downregulation, we next analyzed rIFG modulation during NF (Figure 3). While we did not find any rIFG activation in the VS downregulation and rIFG-VS negative connectivity upregulation groups in comparison to their respective control groups, we found nominally significant lower rIFG activation during NF in the rIFG upregulation group in comparison to control in 6 of 7 runs. Importantly, this is contrary to the intended direction of modulation.

**Figure 3.**
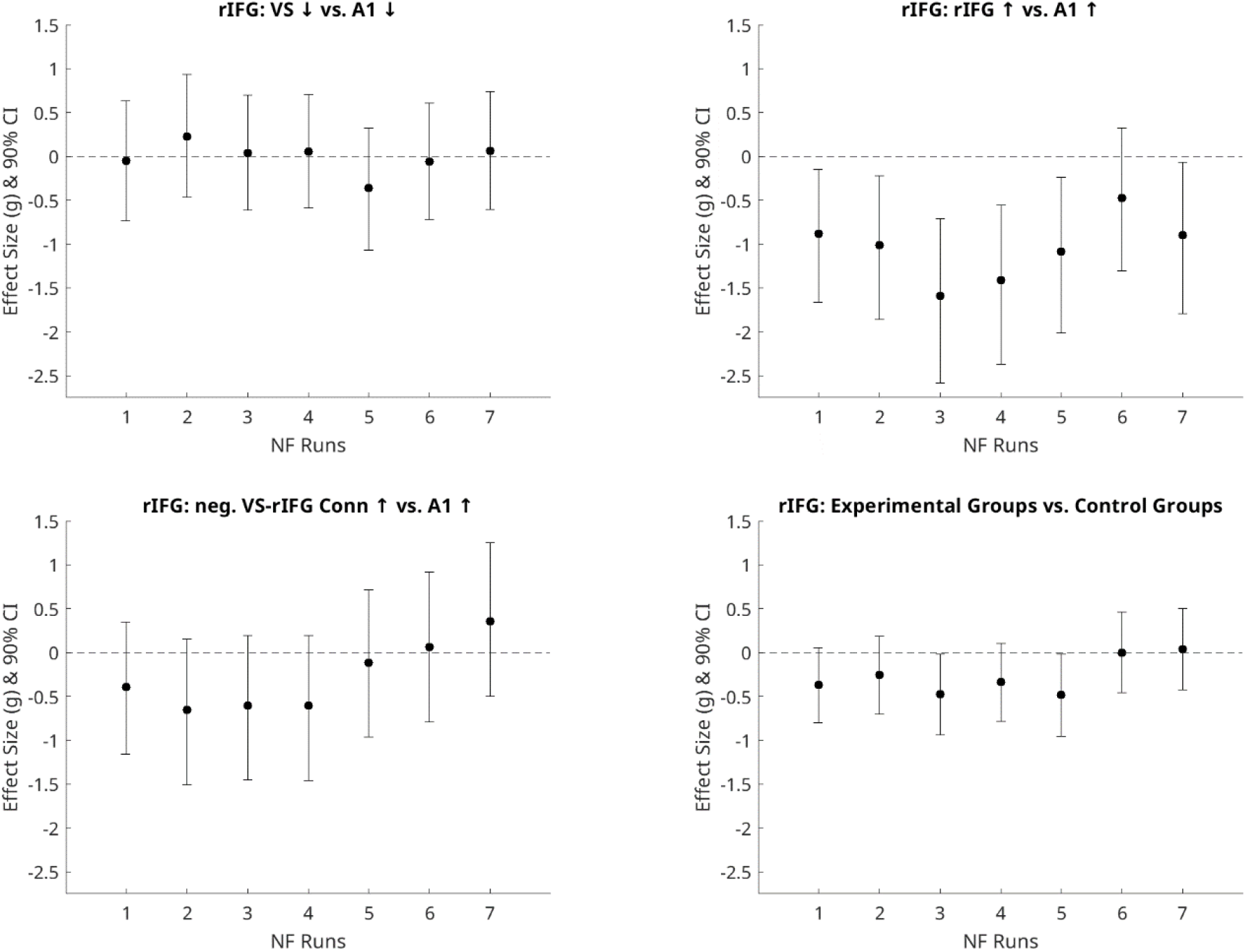
Group comparisons of NF effects in the rIFG. Effect sizes (Hedges’ g) with 90% confidence intervals for group comparisons of rIFG activation with two-sample t-tests across NF runs. Experimental groups are compared with the respective up- or downregulation control groups. In the lower right panel the combined comparison of the experimental groups (VS down / rIFG up / neg. VS-rIFG Conn up) versus the control groups (A1 up / A1 up) is shown. Positive values indicate a higher activation in the rIFG in the experimental groups during NF phases. Please note that the limits of the CI are equivalent to a one-sided uncorrected significance threshold of p<0.05. Exact test statistics for all analyses are provided in the Supplementary Material (Tables S5– S8)

**Figure 4.**
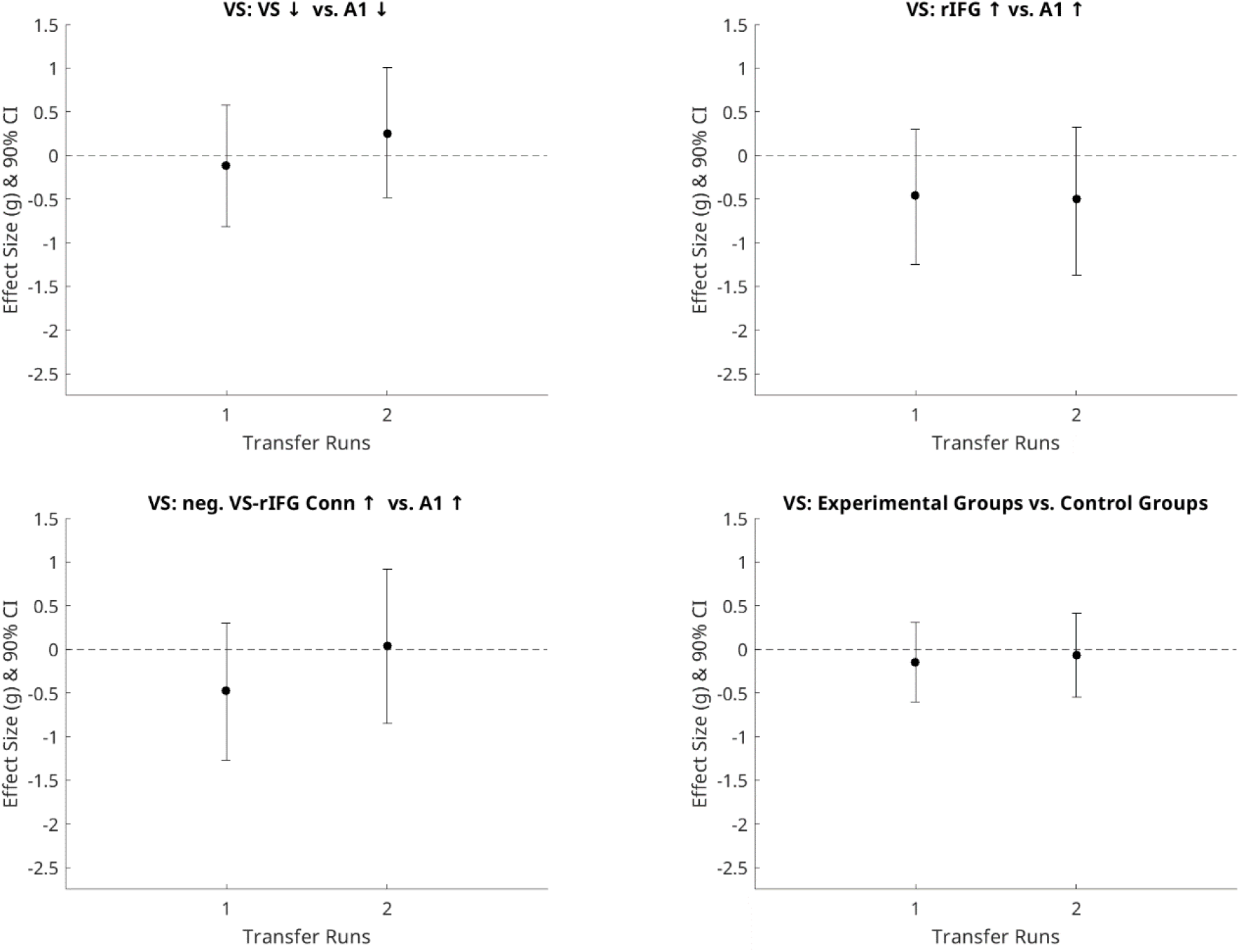
Group comparisons of VS activation in the transfer phase. Effect sizes (Hedges’ g) with 90% confidence intervals for group comparisons of VS activation with two-sample t-tests during transfer runs runs. Experimental groups are compared with the respective up- or downregulation control groups. In the lower right panel the combined comparison of the experimental groups (VS down / rIFG up / neg. VS-rIFG Conn up) versus the control groups (A1 up / A1 up) is shown. Negative values indicate a lower activation in the VS in the experimental groups during NF phases. Exact test statistics for all analyses are provided in the Supplementary Material (Tables S9–S12)

### VS Modulation During Transfer Runs

When changes in VS activation in the first and second transfer runs (which were the third runs of the first and third fMRI session, respectively) were analyzed, no evidence of modulation by NF training in the experimental groups was found. There is also no evidence for a training effect in the experimental groups, which should be visible in transfer run 2.

## Discussion

In this study we investigated the effectiveness of rtfMRI NF for downregulating ventral striatal cue reactivity and enhancing frontal control processes as a potential treatment approach for AUD in a single-blind randomized controlled trial. While we did not find any evidence for a direct downregulation of ventral striatal cue reactivity by direct VS feedback, VS activation was found to be lower in the rIFG upregulation group. rIFG activation was, however, also lower in this group, speaking against a successful modulation of the intended target network. Overall, the study faced several challenges and has clear limitations; we consider it important to nonetheless report our insights, learnings and conclusions in order to inform future work on the application of rtfMRI NF in AUD.

### General Methodological Challenges

The central challenge of targeting specific brain processes lies in their inherent interconnectedness (Emmert et al., 2016). Neural processes such as cue reactivity and reward processing do not operate in isolation but rely on overlapping brain regions and functional networks. For example, the ventral striatum contributes to both processes through shared activations and connectivity patterns making it difficult to modulate one process without affecting the other. One potential way to address this challenge is to target higher-order regulatory regions. The rIFG has been consistently implicated in inhibitory control and top-down regulation of motivational and affective responses, suggesting that modulating this region may influence multiple reward-related processes simultaneously (Koob & Volkow, 2016; Naqvi & Morgenstern, 2015; Schacht et al., 2013).

A growing body of work demonstrates that real-time fMRI NF targeting the ventral striatum, including the NAcc, is technically feasible and can reliably yield volitional upregulation of this reward-related region (Greer et al., 2014; Kirsch et al., 2016; Singer et al., 2023). Evidence for successful downregulation, in contrast, is sparse and inconsistent, suggesting that VS NF may be intrinsically better suited for enhancing rather than suppressing reward-related responses. The difficulty of down-regulating brain activation in regions involved in reward or salience processing might stem from both neurobiological and psychological mechanisms (Heatherton & Wagner, 2011). While increasing activation often aligns with natural cognitive or emotional engagement (e.g., focusing attention or imagining a rewarding stimulus) (Sitaram et al., 2017), reducing activation would require inhibitory control, distraction, or detachment, which are cognitively more demanding and often less intuitive (Diamond, 2013). Further, it requires not just the absence of engagement but the active suppression of internal responses, often without external cues or immediate reinforcement as the feedback of the rtfMRI NF also has a built in delay by design. Particularly in the absence of clear instructions or strategies, this task can be quite challenging and only a very limited number of studies have reported investigating it (Tursic et al., 2020).

From a translational perspective, these findings align with theoretical accounts proposing that strengthening the responsivity to non-drug rewards could help counteract the motivational dominance of substance-related cues in AUD rather than down regulating the response to the substance-related cues (Chung et al., 2023; Li et al., 2018).

### Study-Specific Challenges

A major limitation of our specific experimental design is that successful down-regulation of VS activity itself can elicit a VS reward response to the perceived success (Pereira et al., 2023; Skottnik et al., 2019). Because we delivered feedback continuously, even a successful regulation could create an oscillation of alternating down and up responses. One of the central recommendations for future studies therefore would be to provide VS feedback intermittently after regulation phases, rather than continuously, to minimize this counter-regulatory effect and separate target regulation and reward processing of successful and unsuccessful task performance.

In our study, patients were not informed about the specific neural process targeted for regulation. Instead, they were instructed only to ‘regulate the displayed brain process’ in the specified direction. No specific strategies for modulation were provided. This absence of guidance, combined with the delayed assessment, likely constrained their ability to engage effectively with the task and to develop a clear understanding of the NF mechanism. In the upregulation groups, this lack of transparency may have led to additional confusion, as the instruction to upregulate a brain process in the presence of an alcohol cue could be perceived as counterintuitive.

Extending this line of reasoning, it is important to acknowledge that cue reactivity is not uniformly present across individuals with addiction, as substantial inter-individual variability has been documented in both subjective and neural responses to substance-related cues (Bach et al., 2022; Schacht et al., 2013).Consequently, it cannot be assumed that all participants in our sample exhibited cue-induced activation of the targeted processes. Because no baseline assessment of cue reactivity was conducted prior to the intervention, we are unable to determine whether participants showed measurable cue responsiveness at study entry. We also did not observe any significant group differences in craving induced by the MRI procedure (For details, see Supplementary Table S19 and S20).

We also did not observe a visible training effect across the seven NF runs over the three scanning days. While successful NF training is typically expected to produce at least a gradual trend toward improved regulation across runs, no such pattern was evident in the present data. This finding may reflect the inter-individual variability in rtfMRI NF learning, where a proportion of participants fail to acquire reliable self-regulation of the feedback signal and are therefore often described as non-responders (Emmert et al., 2016; Haugg et al., 2020; Sitaram et al., 2017).

A particularly challenging condition was the connectivity-based NF group, which was instructed to produce a higher negative VS-rIFG correlation. The thermometer provided positive feedback only when VS activity decreased while rIFG activity increased - a strategy that may be effective in down-regulating cue reactivity. However, this task is obviously very difficult. If either region was modulated in the unintended direction, the feedback was negative. As a result, participants might have been unable to determine from the feedback which component of the regulation attempt was unsuccessful.

Another major challenge in this study was that recruitment targets for the planned sample size could not be reached. Unlike many NF trials conducted in convenience samples or highly selected volunteers, this study involved real patients undergoing routine clinical treatment, which substantially increases ecological validity but also places limits on recruitment feasibility. Despite major efforts and an extension of the recruitment period above the originally planned study phase, we were not able to recruit the originally planned full sample of N=100 patients with AUD. In our complex design with 5 groups even the still substantial number of N=65 included patients leaves each group small and underpowered. Retrospectively, a more focused and reduced design would have been preferable. In addition, the participants faced considerable demands - including comprehensive diagnostics, extensive questionnaires, and three MRI session - on top of their treatment as usual. Moreover, enrollment occurred shortly after inpatient admission, during early abstinence and a period often marked by emotional instability, further contributing to reduced willingness or ability to participate. Despite these constraints, session-level retention was high. Very few participants discontinued once training began, suggesting that the intervention was generally well tolerated and perceived as meaningful, even given the inherent difficulty of the NF task. In contrast, retention at follow-up was limited. Although three monthly telephone assessments were planned, only 30 of the 65 participants provided data despite multiple contact attempt, which is consistent with the absence of a successful manipulation of the targeted neural process and therefore with the lack of an expected intervention effect (For details, see Supplementary table S20). Nonetheless, the fact that this trial was conducted in a naturalistic clinical setting with AUD patients enhances its translational relevance and underscores the practical challenges associated with implementing NF interventions in real-world treatment contexts.

### Implications and Future Directions

Recent research in AUD has increasingly focused on methodological optimization and translational integration of rtfMRI NF. Current studies systematically compare single-region-of-interest approaches with multi-ROI and multivariate classifier-based feedback, as well as continuous versus intermittent feedback schedules, to improve target engagement and learning efficacy (Fede et al., 2023). In parallel, rtfMRI NF is progressively embedded within established psychotherapeutic frameworks, such as mindfulness-based relapse prevention, to enhance transfer to everyday self-regulation and relapse prevention (Weiss et al., 2020). However, despite evidence that modulation of prefrontal–striatal circuitry is feasible, recent systematic reviews consistently highlight that robust and sustained clinical effects - particularly regarding craving and alcohol consumption - remain inconsistent (Karch et al., 2022; Murphy et al., 2024).

To enhance NF efficacy in demanding training contexts, combining NF with structured psychotherapeutic interventions appears promising. Integrating cognitive behavioral therapy (CBT) between sessions and reinforcing these strategies during NF may facilitate learning and sustained engagement. Patients can be encouraged to actively implement individualized regulation strategies, with NF serving as a real-time validation mechanism that provides immediate, tangible feedback on strategy effectiveness. This process may strengthen self-efficacy and increase motivation to apply these skills outside the scanner (Fede et al., 2020; MacDuffie et al., 2018). Complementary approaches include mindfulness-based techniques aimed at enhancing attentional control and self-regulation, thereby supporting more effective modulation of the NF signal (Tang et al., 2015). In clinical populations, particularly in addiction, emphasizing long-term goals may preferentially recruit prefrontal control networks and reduce the salience of immediate, disorder-relevant rewards (Koob & Volkow, 2016). Additionally, interoceptive focus represents a promising strategy for fine-grained self-regulation. Participants may be instructed to attend to subtle bodily sensations and deliberately release micro-tensions (e.g., in the jaw or shoulders) while observing corresponding changes in the feedback signal (Sugawara et al., 2024). This approach may facilitate a more precise mapping between internal bodily states and neural activity, thereby improving NF modulation. Finally, affect labeling—i.e., the verbal or mental articulation of one’s current emotional state (e.g., “I notice tension and craving; they rise and then subside”) - has been shown to attenuate limbic reactivity via recruitment of prefrontal regulatory networks (Torre & Lieberman, 2018). This mechanism may help reduce disorder-relevant cue reactivity and further support voluntary control over the NF signal.

Taken together, our study especially demonstrates the challenges when targeting VS cue reactivity in a rtfMRI NF study, but does not finally answer the question whether this approach could work if the mentioned modifications are taken into account.

### Outlook

Our study highlights key challenges for the modulation of targeted brain regions with rtfMRI NF in patients with alcohol use disorder. Because cue reactivity, reward processing, and cognitive control rely on overlapping networks and partly entangled processes, selectively downregulating ventral striatal activation proved difficult. Continuous feedback was likely a specific problem in our study and using intermittent feedback may help mitigate this issue. In addition, the lack of explicit strategy guidance and transparency about the targeted process likely limited patients’ engagement, particularly in the rIFG condition. Future work should refine feedback timing, provide clearer instructions and regulation strategies, and further clarify how neural modulation may translate into meaningful clinical change.

## Statements

## Acknowledgement

ChatGPT (OpenAI) was used exclusively for language editing and proofreading. All scientific content, interpretations and conclusions were created and verified by the authors.

## Statement of Ethics

This study was performed in accordance with the Declaration of Helsinki. This human study was approved by Ethics Committee of the Medical Faculty Mannheim at the University of Heidelberg, Germany - approval: 2015-613N-MA. The study’s clinical trial registration number is DRKS00010253 registered with German Clinical Trials Register (https://drks.de/). Participant registration took place from March 2016 to October 2020. All adult participants provided written informed consent to participate in this study.

## Conflict of Interest Statement

The authors declare that the research was conducted in the absence of any commercial or financial relationships that could be construed as a potential conflict of interest.

## Funding

The study was supported by the European Union’s Horizon 2020 research and innovation program: 668863-SyBil-AA and the Deutsche Forschungsgemeinschaft (DFG, German Research Foundation): Project-ID 402170461 - TRR 265 (Spanagel et al., 2024)

## Author Contributions

Patrick Halli: Data curation, Formal analysis, Methodology, Investigation, Software, Visualization, Writing – original draft, Writing – review & editing.

Franziska Weiss: Validation, Investigation, Writing – review & editing

Sarah Gerhardt: Data curation, Investigation, Writing – review & editing

Jingying Zhang: Validation, Investigation, Writing – review & editing

Wolfgang H. Sommer: Funding acquisition, Resources, Writing – review & editing

Falk Kiefer: Conceptualization, Funding acquisition, Resources, Writing – review & editing

Peter Kirsch: Conceptualization, Funding acquisition, Methodology, Project administration, Resources, Supervision, Writing – review & editing.

Martin Fungisai Gerchen: Conceptualization, Funding Acquisition, Methodology, Investigation, Formal analysis, Data curation, Project administration, Software, Supervision, Visualization, Writing – original draft, Writing – review & editing.

## Data Availability Statement

Due to ethical and legal constraints associated with clinical patient data, the datasets generated and analyzed in this study cannot be made publicly available. Data may be shared upon reasonable request for scientific purposes.

## Notes

### Competing Interest Statement

The authors have declared no competing interest.

### Clinical Trial

DRKS00010253

