## Supplement for "Real-Time fMRI Neurofeedback Targeting Cue Reactivity in Alcohol Use Disorder: Challenges and Insights from a Randomized Controlled Trial"

Tables S1–S8. Results of one-sample t-tests across neurofeedback runs.

Note. t = t-statistic; df = degrees of freedom; g = Hedges' g; CI = confidence interval. All tests are two-tailed. NF1–NF7 denote neurofeedback runs.

*Table S1 VS: VS ↓ vs. A1 ↓ (Figure 2 upper left)*

| Run | t | df | p | g | 90% CI for g |
|---|---|---|---|---|---|
| NF1 | -0.958 | 24 | .347 | -0.386 | [-1.085, 0.296] |
| NF2 | -0.505 | 22 | .619 | -0.207 | [-0.911, 0.488] |
| NF3 | -0.863 | 23 | .397 | -0.334 | [-1.004, 0.321] |
| NF4 | 0.243 | 25 | .810 | 0.092 | [-0.550, 0.739] |
| NF5 | -0.896 | 21 | .381 | -0.361 | [-1.064, 0.324] |
| NF6 | -0.359 | 22 | .723 | -0.140 | [-0.809, 0.522] |
| NF7 | -0.896 | 23 | .380 | -0.353 | [-1.038, 0.315] |

*Table S2 VS: rIFG ↑ vs A1 ↑ (Figure 2 upper right)*

| Run | t | df | p | g | 90% CI for g |
|---|---|---|---|---|---|
| NF1 | -2.516 | 19 | .021 | -1.055 | [-1.861, -0.311] |
| NF2 | -2.057 | 18 | .054 | -0.917 | [-1.754, -0.136] |
| NF3 | -1.982 | 16 | .065 | -0.920 | [-1.800, -0.103] |
| NF4 | -2.184 | 16 | .044 | -1.013 | [-1.906, -0.191] |
| NF5 | -2.214 | 15 | .043 | -1.048 | [-1.972, -0.204] |
| NF6 | -2.247 | 15 | .040 | -1.041 | [-1.947, -0.214] |
| NF7 | -1.557 | 14 | .142 | -0.728 | [-1.604, 0.090] |

*Table S3 VS: VS: neg. VS-rIFG Conn ↑ vs. A1 ↑(Figure 2 lower left)*

| Run | t | df | p | g | 90% CI for g |
|---|---|---|---|---|---|
| NF1 | -2.161 | 16 | .046 | -0.927 | [-1.752, -0.167] |
| NF2 | -1.070 | 14 | .303 | -0.494 | [-1.332, 0.305] |
| NF3 | -0.476 | 14 | .642 | -0.219 | [-1.032, 0.577] |
| NF4 | -0.292 | 14 | .775 | -0.134 | [-0.942, 0.663] |
| NF5 | -0.511 | 13 | .618 | -0.244 | [-1.095, 0.585] |
| NF6 | 0.474 | 12 | .644 | 0.231 | [-0.619, 1.102] |
| NF7 | 0.147 | 12 | .886 | 0.072 | [-0.791, 0.942] |

*Table S4 VS: Experimental Groups vs. Control Groups (Figure 2 lower right)*

| Run | t | df | p | g | 90% CI for g |
|---|---|---|---|---|---|
| NF1 | -2.216 | 58 | .031 | -0.565 | [-1.005, -0.136] |
| NF2 | -1.338 | 54 | .187 | -0.354 | [-0.802, 0.088] |
| NF3 | -1.650 | 53 | .105 | -0.445 | [-0.906, 0.006] |
| NF4 | -1.207 | 55 | .233 | -0.319 | [-0.765, 0.121] |
| NF5 | -1.659 | 49 | .103 | -0.460 | [-0.934, 0.005] |
| NF6 | -0.844 | 49 | .403 | -0.233 | [-0.698, 0.227] |
| NF7 | -0.877 | 49 | .385 | -0.243 | [-0.709, 0.219] |

*Table S5 rIFG: rIFG: VS ↓ vs. A1 ↓(Figure 3 upper left)*

| Run | t | df | p | g | 90% CI for g |
|---|---|---|---|---|---|
| NF1 | -0.129 | 24 | .899 | -0.052 | [-0.737, 0.631] |
| NF2 | 0.557 | 22 | .583 | 0.228 | [-0.466, 0.933] |
| NF3 | 0.110 | 23 | .913 | 0.043 | [-0.614, 0.701] |
| NF4 | 0.150 | 25 | .882 | 0.057 | [-0.586, 0.703] |
| NF5 | -0.899 | 21 | .379 | -0.362 | [-1.066, 0.323] |
| NF6 | -0.144 | 22 | .887 | -0.056 | [-0.722, 0.607] |
| NF7 | 0.159 | 23 | .875 | 0.063 | [-0.607, 0.735] |

*Table S6 rIFG: rIFG: rIFG ↑ vs. A1 ↑( Figure 3 upper right)*

| Run | t | df | p | g | 90% CI for g |
|---|---|---|---|---|---|
| NF1 | -2.104 | 19 | .049 | -0.882 | [-1.666, -0.149] |
| NF2 | -2.271 | 18 | .036 | -1.012 | [-1.861, -0.226] |
| NF3 | -3.426 | 16 | .003 | -1.589 | [-2.582, -0.712] |
| NF4 | -3.040 | 16 | .008 | -1.410 | [-2.369, -0.553] |
| NF5 | -2.286 | 15 | .037 | -1.083 | [-2.012, -0.235] |
| NF6 | -1.023 | 15 | .323 | -0.474 | [-1.307, 0.325] |
| NF7 | -1.916 | 14 | .076 | -0.896 | [-1.796, -0.068] |

*Table S7 rIFG: rIFG: neg. VS-rIFG Conn ↑ vs. A1 ↑(Figure 3 lower left)*

| Run | t | df | p | g | 90% CI for g |
|---|---|---|---|---|---|
| NF1 | -0.915 | 16 | .374 | -0.393 | [-1.156, 0.344] |
| NF2 | -1.416 | 14 | .179 | -0.654 | [-1.510, 0.151] |
| NF3 | -1.314 | 14 | .210 | -0.605 | [-1.454, 0.196] |
| NF4 | -1.318 | 14 | .209 | -0.607 | [-1.457, 0.194] |
| NF5 | -0.244 | 13 | .811 | -0.117 | [-0.959, 0.715] |
| NF6 | 0.126 | 12 | .902 | 0.061 | [-0.793, 0.921] |
| NF7 | 0.728 | 12 | .480 | 0.359 | [-0.500, 1.251] |

*Table S8 rIFG: Experimental Groups vs. Control Groups (Figure 3 lower right)*

| Run | t | df | p | g | 90% CI for g |
|---|---|---|---|---|---|
| NF1 | -1.450 | 58 | .152 | -0.370 | [-0.802, 0.056] |
| NF2 | -0.968 | 54 | .337 | -0.256 | [-0.701, 0.185] |
| NF3 | -1.755 | 53 | .085 | -0.474 | [-0.935, -0.022] |
| NF4 | -1.277 | 55 | .207 | -0.337 | [-0.784, 0.103] |
| NF5 | -1.741 | 49 | .088 | -0.482 | [-0.957, -0.018] |
| NF6 | 0.005 | 49 | .996 | 0.001 | [-0.459, 0.462] |
| NF7 | 0.140 | 49 | .889 | 0.039 | [-0.423, 0.501] |

Tables S9–S12. Results of one-sample t-tests across neurofeedback transfer runs.

Note. t = t-statistic; df = degrees of freedom; g = Hedges' g; CI = confidence interval. All tests are two-tailed. Transfer 1-2 denote neurofeedback transfer runs.

*Table S9 VS Transfer: VS ↓ vs. A1 ↓ (Figure 4 upper left)*

| Run | t | df | p | g | 90% CI for g |
|---|---|---|---|---|---|
| Transfer1 | -0.295 | 20 | .771 | -0.120 | [-0.820, 0.574] |
| Transfer2 | 0.586 | 19 | .565 | 0.253 | [-0.483, 1.004] |

*Table S10 VS Transfer: rIFG ↑ vs A1 ↑ (Figure 4 upper right)*

| Run | t | df | p | g | 90% CI for g |
|---|---|---|---|---|---|
| Transfer1 | -1.033 | 17 | .316 | -0.458 | [-1.248, 0.302] |
| Transfer2 | -1.049 | 14 | .312 | -0.502 | [-1.369, 0.327] |

*Table S11 VS Transfer: neg. VS-rIFG Conn ↑ vs. A1 ↑(Figure 4 lower left)*

| Run | t | df | p | g | 90% CI for g |
|---|---|---|---|---|---|
| Transfer1 | -1.060 | 15 | .306 | -0.471 | [-1.272, 0.296] |
| Transfer2 | 0.069 | 12 | .946 | 0.035 | [-0.849, 0.921] |

*Table S12 VS Transfer: Experimental Groups vs. Control Groups (Figure 4 lower right)*

| Run | t | df | p | g | 90% CI for g |
|---|---|---|---|---|---|
| Transfer1 | -0.529 | 52 | .599 | -0.145 | [-0.604, 0.311] |
| Transfer2 | -0.238 | 46 | .813 | -0.068 | [-0.549, 0.411] |

One Sample t-Tests (wird noch schöngemacht, wenn final)

*Table S13 VS: Neurofeedback Runs*

| Group | Run | t | df | p | g | 90% CI for g |
|---|---|---|---|---|---|---|
| VS ↓ | NF1 | 0.747 | 10 | .472 | 0.208 | [-0.256, 0.672] |
| VS ↓ | NF2 | 0.753 | 9 | .471 | 0.218 | [-0.264, 0.700] |
| VS ↓ | NF3 | -0.026 | 12 | .980 | -0.007 | [-0.434, 0.420] |
| VS ↓ | NF4 | 1.030 | 12 | .323 | 0.267 | [-0.168, 0.703] |
| VS ↓ | NF5 | 0.860 | 10 | .410 | 0.239 | [-0.226, 0.705] |
| VS ↓ | NF6 | 0.218 | 11 | .831 | 0.059 | [-0.384, 0.501] |
| VS ↓ | NF7 | 0.548 | 11 | .595 | 0.147 | [-0.297, 0.592] |
| rIFG ↑ | NF1 | -0.607 | 13 | .554 | -0.153 | [-0.569, 0.264] |
| rIFG ↑ | NF2 | -1.160 | 13 | .267 | -0.292 | [-0.715, 0.132] |
| rIFG ↑ | NF3 | -0.735 | 11 | .478 | -0.197 | [-0.644, 0.249] |
| rIFG ↑ | NF4 | -1.071 | 11 | .307 | -0.288 | [-0.740, 0.165] |
| rIFG ↑ | NF5 | -0.880 | 10 | .400 | -0.245 | [-0.711, 0.221] |
| rIFG ↑ | NF6 | -0.896 | 10 | .391 | -0.249 | [-0.716, 0.217] |
| rIFG ↑ | NF7 | -1.141 | 9 | .283 | -0.330 | [-0.821, 0.161] |
| Neg. VS–rIFG Conn ↑ | NF1 | -0.689 | 10 | .507 | -0.192 | [-0.654, 0.271] |
| Neg. VS–rIFG Conn ↑ | NF2 | -0.144 | 9 | .889 | -0.041 | [-0.517, 0.434] |
| Neg. VS–rIFG Conn ↑ | NF3 | 1.259 | 9 | .240 | 0.364 | [-0.130, 0.858] |
| Neg. VS–rIFG Conn ↑ | NF4 | 0.984 | 9 | .351 | 0.285 | [-0.202, 0.772] |
| Neg. VS–rIFG Conn ↑ | NF5 | -0.049 | 8 | .962 | -0.015 | [-0.510, 0.481] |
| Neg. VS–rIFG Conn ↑ | NF6 | 2.528 | 7 | .039 | 0.795 | [0.183, 1.406] |
| Neg. VS–rIFG Conn ↑ | NF7 | 0.166 | 7 | .873 | 0.052 | [-0.465, 0.570] |
| A1 ↑ | NF1 | 2.746 | 8 | .025 | 0.827 | [0.237, 1.417] |
| A1 ↑ | NF2 | 2.248 | 7 | .059 | 0.707 | [0.114, 1.300] |
| A1 ↑ | NF3 | 1.862 | 7 | .105 | 0.585 | [0.015, 1.156] |
| A1 ↑ | NF4 | 1.965 | 7 | .090 | 0.617 | [0.041, 1.193] |
| A1 ↑ | NF5 | 1.512 | 7 | .174 | 0.475 | [-0.077, 1.028] |
| A1 ↑ | NF6 | 2.238 | 7 | .060 | 0.703 | [0.111, 1.296] |
| A1 ↑ | NF7 | -0.227 | 7 | .827 | -0.071 | [-0.589, 0.446] |
| A1 ↓ | NF1 | 2.529 | 16 | .022 | 0.584 | [0.170, 0.998] |
| A1 ↓ | NF2 | 0.869 | 15 | .399 | 0.206 | [-0.189, 0.601] |
| A1 ↓ | NF3 | 2.157 | 13 | .050 | 0.543 | [0.096, 0.989] |
| A1 ↓ | NF4 | 1.679 | 15 | .114 | 0.398 | [-0.009, 0.806] |
| A1 ↓ | NF5 | 2.090 | 13 | .057 | 0.526 | [0.081, 0.971] |
| A1 ↓ | NF6 | 1.276 | 13 | .224 | 0.321 | [-0.105, 0.747] |
| A1 ↓ | NF7 | 1.364 | 14 | .194 | 0.333 | [-0.081, 0.747] |

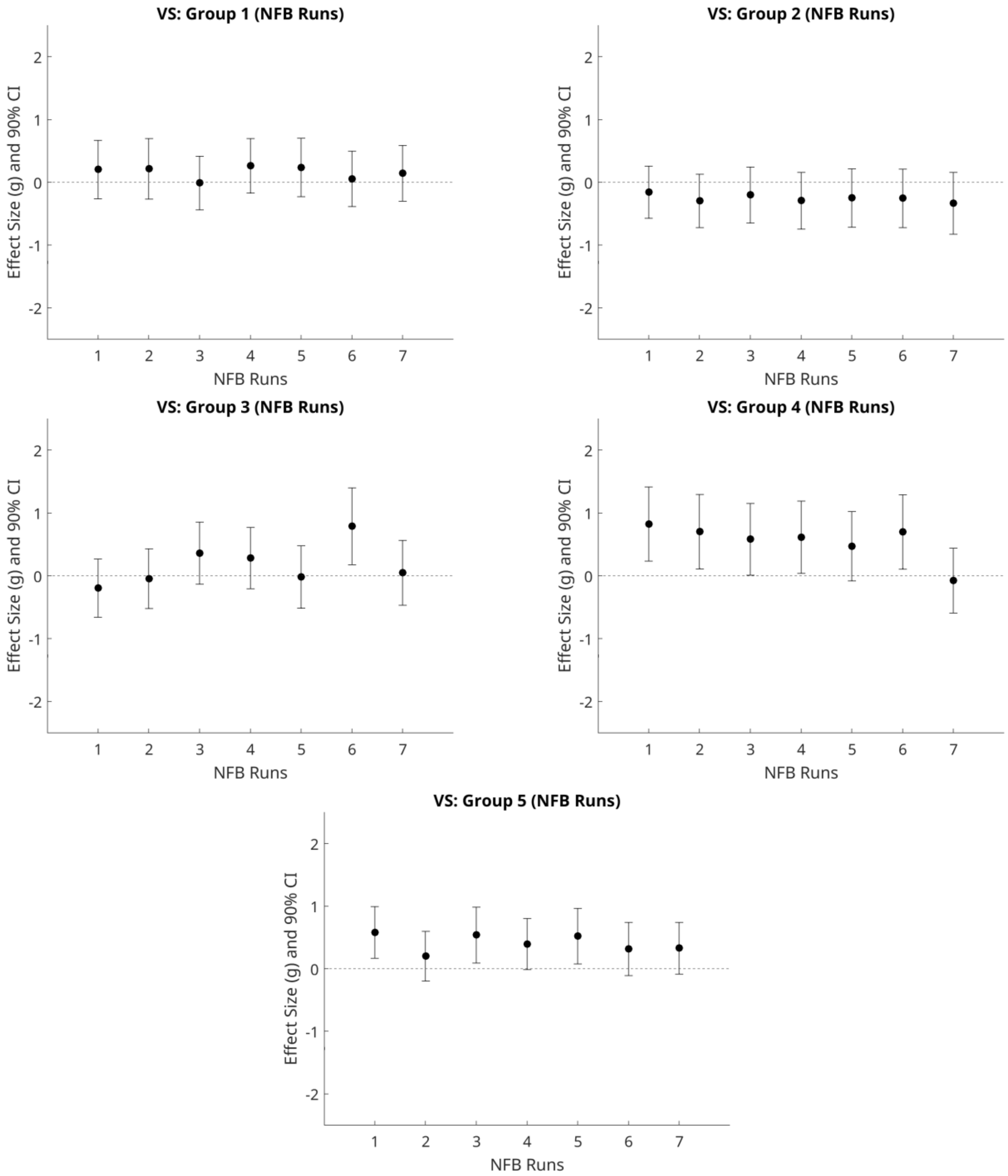


**Figure S1. Within-group effects on basic target process modulation: VS**

Points represent Hedges' g for each neurofeedback (NFB) run (1–7), with error bars indicating 90% confidence intervals. The dashed horizontal line denotes zero effect. Panels display results separately for each group: Group 1 (VS ↓), Group 2 (rIFG ↑), Group 3 (negative VS–rIFG connectivity ↑), Group 4 (A1 ↑), and Group 5 (A1 ↓).

*Table S14 rIFG: Neurofeedback Runs*

| Group | Run | t | df | p | g | 90% CI for g |
|---|---|---|---|---|---|---|
| VS ↓ | NF1 | 5.751 | 10 | .000 | 1.600 | [0.876, 2.325] |
| VS ↓ | NF2 | 5.463 | 9 | .000 | 1.579 | [0.829, 2.330] |
| VS ↓ | NF3 | 4.706 | 12 | .001 | 1.222 | [0.641, 1.803] |
| VS ↓ | NF4 | 5.017 | 12 | .000 | 1.303 | [0.703, 1.902] |
| VS ↓ | NF5 | 3.367 | 10 | .007 | 0.937 | [0.374, 1.501] |
| VS ↓ | NF6 | 3.398 | 11 | .006 | 0.912 | [0.375, 1.450] |
| VS ↓ | NF7 | 2.990 | 11 | .012 | 0.803 | [0.285, 1.320] |
| rIFG ↑ | NF1 | 3.641 | 13 | .003 | 0.916 | [0.414, 1.418] |
| rIFG ↑ | NF2 | 4.468 | 13 | .001 | 1.124 | [0.582, 1.665] |
| rIFG ↑ | NF3 | 2.448 | 11 | .032 | 0.657 | [0.164, 1.151] |
| rIFG ↑ | NF4 | 1.953 | 11 | .077 | 0.524 | [0.049, 1.000] |
| rIFG ↑ | NF5 | 0.803 | 10 | .441 | 0.224 | [-0.241, 0.688] |
| rIFG ↑ | NF6 | 1.869 | 10 | .091 | 0.520 | [0.027, 1.013] |
| rIFG ↑ | NF7 | 0.598 | 9 | .565 | 0.173 | [-0.307, 0.653] |
| Neg. VS–rIFG Conn ↑ | NF1 | 2.455 | 10 | .034 | 0.683 | [0.167, 1.200] |
| Neg. VS–rIFG Conn ↑ | NF2 | 4.394 | 9 | .002 | 1.270 | [0.604, 1.937] |
| Neg. VS–rIFG Conn ↑ | NF3 | 2.947 | 9 | .016 | 0.852 | [0.283, 1.422] |
| Neg. VS–rIFG Conn ↑ | NF4 | 2.787 | 9 | .021 | 0.806 | [0.245, 1.366] |
| Neg. VS–rIFG Conn ↑ | NF5 | 3.011 | 8 | .017 | 0.907 | [0.299, 1.514] |
| Neg. VS–rIFG Conn ↑ | NF6 | 5.429 | 7 | .001 | 1.706 | [0.835, 2.577] |
| Neg. VS–rIFG Conn ↑ | NF7 | 6.513 | 7 | .000 | 2.047 | [1.059, 3.035] |
| A1 ↑ | NF1 | 7.990 | 8 | .000 | 2.406 | [1.350, 3.462] |
| A1 ↑ | NF2 | 13.029 | 7 | .000 | 4.095 | [2.333, 5.856] |
| A1 ↑ | NF3 | 5.996 | 7 | .001 | 1.884 | [0.953, 2.816] |
| A1 ↑ | NF4 | 6.570 | 7 | .000 | 2.065 | [1.071, 3.059] |
| A1 ↑ | NF5 | 6.981 | 7 | .000 | 2.194 | [1.154, 3.234] |
| A1 ↑ | NF6 | 5.687 | 7 | .001 | 1.787 | [0.889, 2.686] |
| A1 ↑ | NF7 | 4.019 | 7 | .005 | 1.263 | [0.530, 1.996] |
| A1 ↓ | NF1 | 6.990 | 16 | .000 | 1.615 | [1.021, 2.208] |
| A1 ↓ | NF2 | 5.226 | 15 | .000 | 1.240 | [0.709, 1.771] |
| A1 ↓ | NF3 | 4.500 | 13 | .001 | 1.132 | [0.589, 1.675] |
| A1 ↓ | NF4 | 3.653 | 15 | .002 | 0.867 | [0.402, 1.332] |
| A1 ↓ | NF5 | 4.270 | 13 | .001 | 1.074 | [0.542, 1.606] |
| A1 ↓ | NF6 | 3.838 | 13 | .002 | 0.966 | [0.454, 1.477] |
| A1 ↓ | NF7 | 1.668 | 14 | .117 | 0.407 | [-0.012, 0.827] |

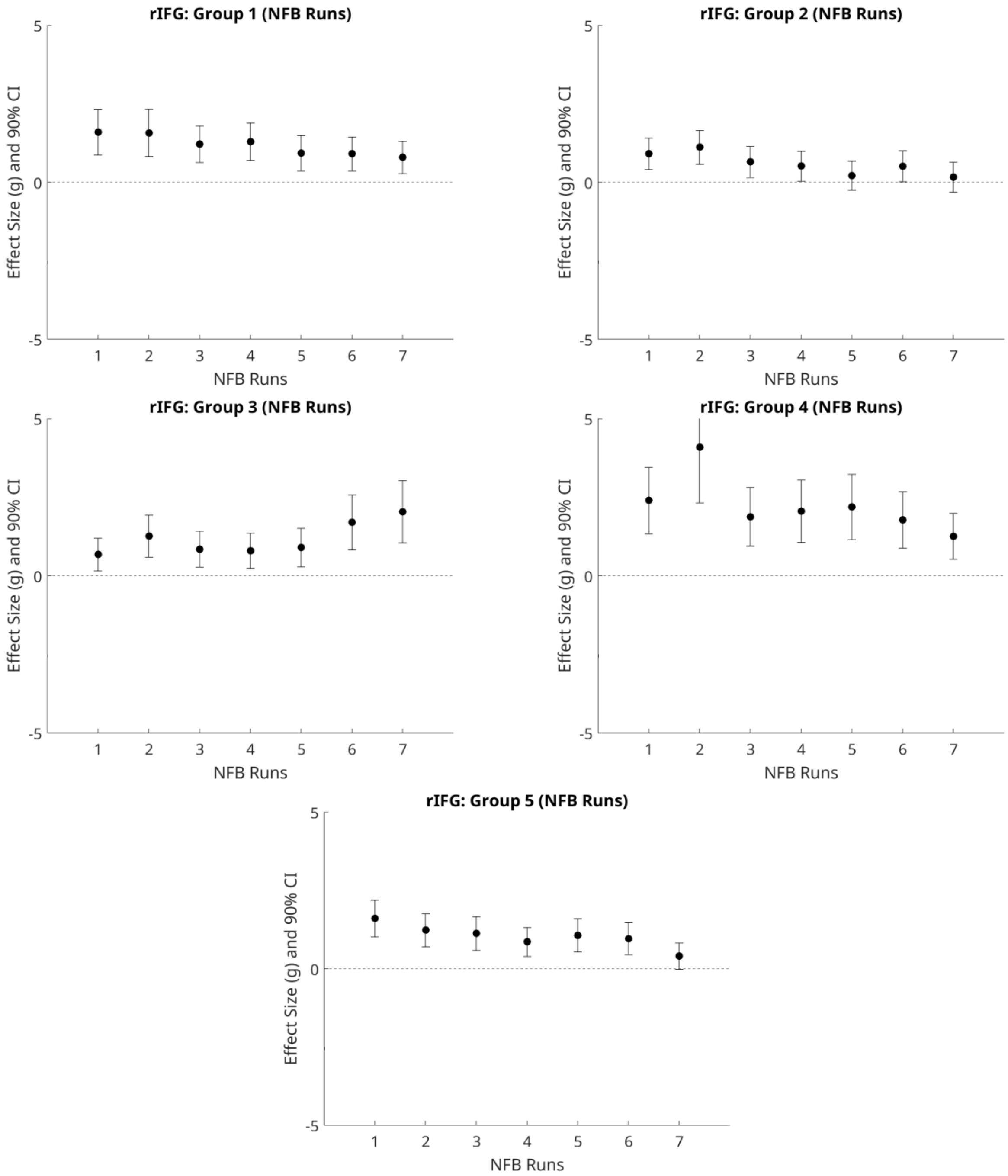


**Figure S2. Within-group effects on basic target process modulation: rIFG**

Points represent Hedges' g for each neurofeedback (NFB) run (1–7), with error bars indicating 90% confidence intervals. The dashed horizontal line denotes zero effect. Panels display results separately for each group: Group 1 (VS ↓), Group 2 (rIFG ↑), Group 3 (negative VS–rIFG connectivity ↑), Group 4 (A1 ↑), and Group 5 (A1 ↓).

*Table S15 A1: Neurofeedback Runs*

| Group | Run | t | df | p | g | 90% CI for g |
|---|---|---|---|---|---|---|
| VS ↓ | NF1 | -0.186 | 10 | .856 | -0.052 | [-0.510, 0.406] |
| VS ↓ | NF2 | -0.145 | 9 | .888 | -0.042 | [-0.518, 0.434] |
| VS ↓ | NF3 | -0.389 | 12 | .704 | -0.101 | [-0.529, 0.327] |
| VS ↓ | NF4 | 0.543 | 12 | .597 | 0.141 | [-0.289, 0.570] |
| VS ↓ | NF5 | -0.296 | 10 | .774 | -0.082 | [-0.541, 0.376] |
| VS ↓ | NF6 | -0.848 | 11 | .414 | -0.228 | [-0.676, 0.221] |
| VS ↓ | NF7 | -0.893 | 11 | .391 | -0.240 | [-0.689, 0.209] |
| rIFG ↑ | NF1 | -2.290 | 13 | .039 | -0.576 | [-1.027, -0.125] |
| rIFG ↑ | NF2 | -2.379 | 13 | .033 | -0.598 | [-1.052, -0.145] |
| rIFG ↑ | NF3 | -4.214 | 11 | .001 | -1.131 | [-1.714, -0.549] |
| rIFG ↑ | NF4 | -2.885 | 11 | .015 | -0.775 | [-1.287, -0.262] |
| rIFG ↑ | NF5 | -3.092 | 10 | .011 | -0.861 | [-1.409, -0.312] |
| rIFG ↑ | NF6 | -2.323 | 10 | .043 | -0.647 | [-1.157, -0.136] |
| rIFG ↑ | NF7 | -4.103 | 9 | .003 | -1.186 | [-1.832, -0.541] |
| Neg. VS–rIFG Conn ↑ | NF1 | -0.734 | 10 | .480 | -0.204 | [-0.668, 0.259] |
| Neg. VS–rIFG Conn ↑ | NF2 | -1.529 | 9 | .161 | -0.442 | [-0.945, 0.061] |
| Neg. VS–rIFG Conn ↑ | NF3 | -0.993 | 9 | .347 | -0.287 | [-0.774, 0.200] |
| Neg. VS–rIFG Conn ↑ | NF4 | -0.707 | 9 | .498 | -0.204 | [-0.686, 0.277] |
| Neg. VS–rIFG Conn ↑ | NF5 | -1.218 | 8 | .258 | -0.367 | [-0.882, 0.148] |
| Neg. VS–rIFG Conn ↑ | NF6 | -0.870 | 7 | .413 | -0.273 | [-0.803, 0.256] |
| Neg. VS–rIFG Conn ↑ | NF7 | -1.573 | 7 | .160 | -0.494 | [-1.050, 0.061] |
| A1 ↑ | NF1 | 2.229 | 8 | .056 | 0.671 | [0.112, 1.231] |
| A1 ↑ | NF2 | 2.617 | 7 | .035 | 0.822 | [0.205, 1.440] |
| A1 ↑ | NF3 | 2.289 | 7 | .056 | 0.719 | [0.124, 1.315] |
| A1 ↑ | NF4 | 0.299 | 7 | .774 | 0.094 | [-0.424, 0.612] |
| A1 ↑ | NF5 | 0.388 | 7 | .710 | 0.122 | [-0.397, 0.641] |
| A1 ↑ | NF6 | 1.196 | 7 | .271 | 0.376 | [-0.164, 0.915] |
| A1 ↑ | NF7 | -1.316 | 7 | .230 | -0.414 | [-0.958, 0.131] |
| A1 ↓ | NF1 | 0.019 | 16 | .985 | 0.004 | [-0.376, 0.384] |
| A1 ↓ | NF2 | -0.285 | 15 | .780 | -0.068 | [-0.458, 0.323] |
| A1 ↓ | NF3 | -0.945 | 13 | .362 | -0.238 | [-0.658, 0.183] |
| A1 ↓ | NF4 | -0.793 | 15 | .440 | -0.188 | [-0.582, 0.206] |
| A1 ↓ | NF5 | -1.057 | 13 | .310 | -0.266 | [-0.688, 0.156] |
| A1 ↓ | NF6 | -1.148 | 13 | .272 | -0.289 | [-0.712, 0.135] |
| A1 ↓ | NF7 | -1.393 | 14 | .185 | -0.340 | [-0.754, 0.074] |

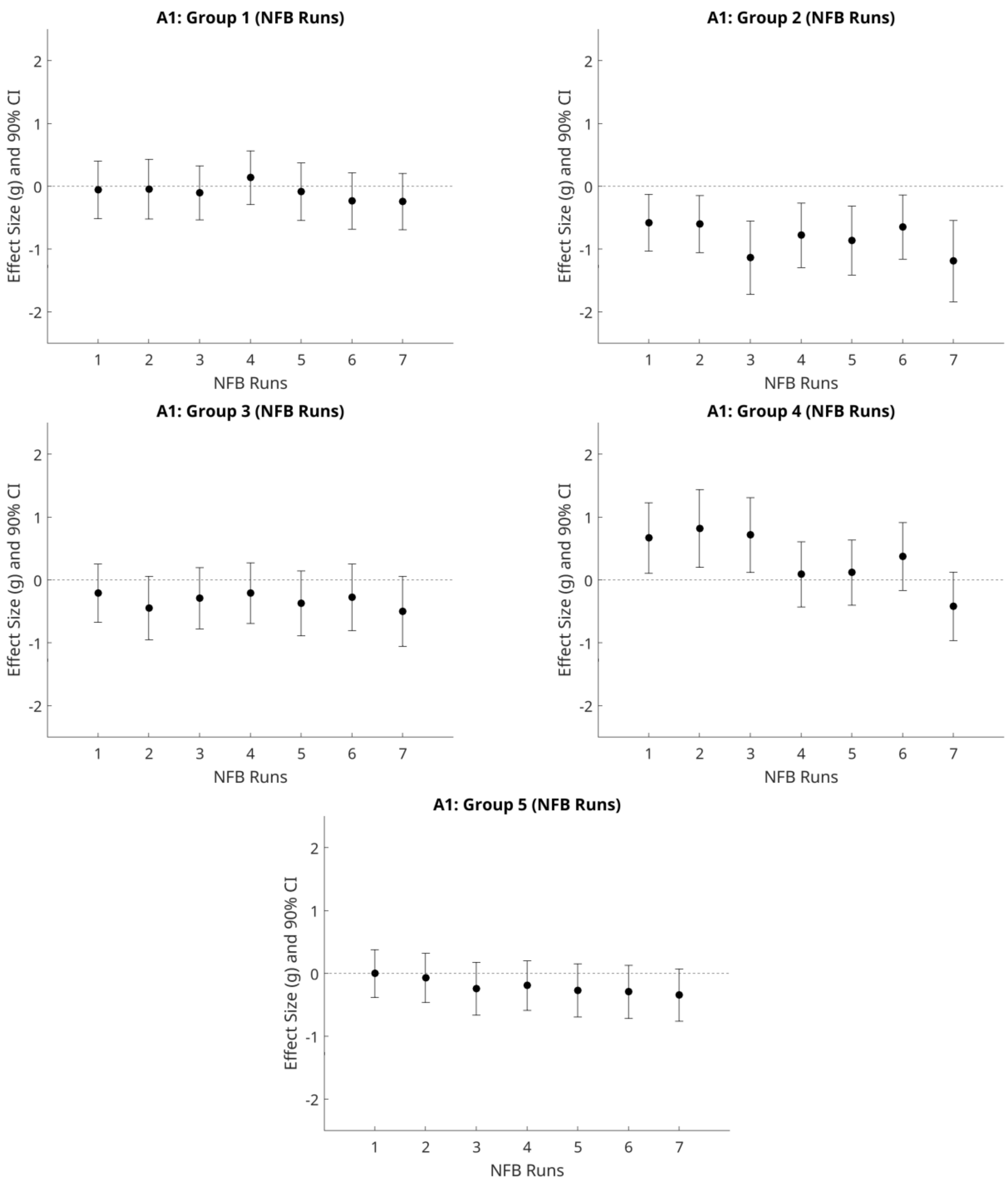


**Figure S3. Within-group effects on basic target process modulation: A1**

Points represent Hedges' g for each neurofeedback (NFB) run (1–7), with error bars indicating 90% confidence intervals. The dashed horizontal line denotes zero effect. Panels display results separately for each group: Group 1 (VS ↓), Group 2 (rIFG ↑), Group 3 (negative VS–rIFG connectivity ↑), Group 4 (A1 ↑), and Group 5 (A1 ↓).

Tables S16–S18. Results of one-sample t-tests for transfer runs across regions of interest and groups.

Note. t = t-statistic; df = degrees of freedom; g = Hedges' g; CI = confidence interval.

*Table S16 VS: Transfer Runs*

| **Group** | **Run** | **t** | **df** | **p** | **g** | **90% CI for g** |
|---|---|---|---|---|---|---|
| VS ↓ | Transfer1 | -1.434 | 10 | .182 | -0.399 | [-0.878, 0.080] |
| VS ↓ | Transfer2 | 1.681 | 8 | .131 | 0.506 | [-0.027, 1.039] |
| rIFG ↑ | Transfer1 | -0.135 | 12 | .895 | -0.035 | [-0.462, 0.392] |
| rIFG ↑ | Transfer2 | -1.337 | 10 | .211 | -0.372 | [-0.848, 0.104] |
| Neg. VS–rIFG Conn ↑ | Transfer1 | -0.465 | 10 | .652 | -0.130 | [-0.590, 0.330] |
| Neg. VS–rIFG Conn ↑ | Transfer2 | 0.199 | 8 | .847 | 0.060 | [-0.436, 0.556] |
| A1 ↑ | Transfer1 | 1.391 | 7 | .207 | 0.437 | [-0.110, 0.984] |
| A1 ↑ | Transfer2 | -0.543 | 6 | .607 | -0.179 | [-0.725, 0.368] |
| A1 ↓ | Transfer1 | -1.301 | 12 | .218 | -0.338 | [-0.779, 0.103] |
| A1 ↓ | Transfer2 | 0.248 | 13 | .808 | 0.062 | [-0.352, 0.477] |

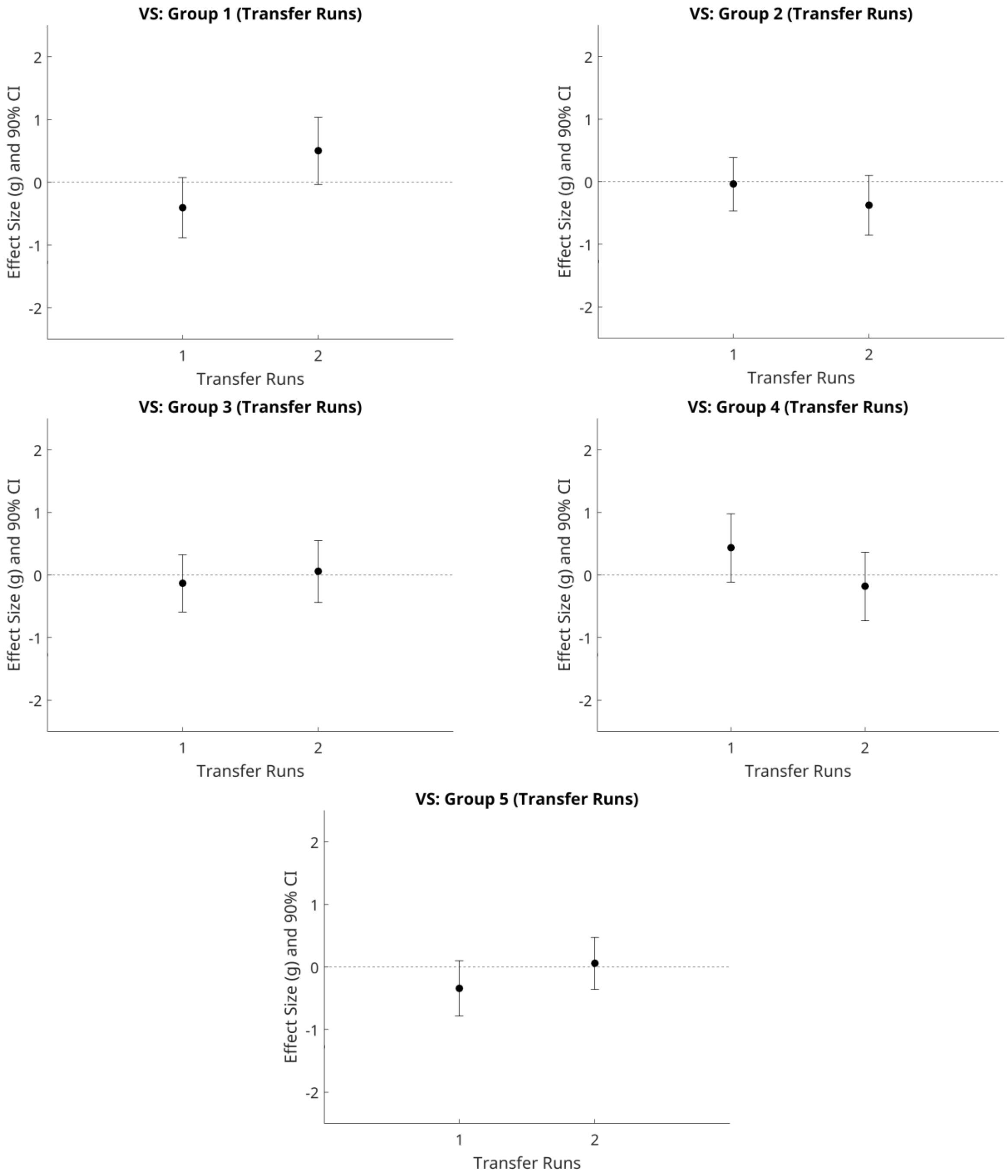


**Figure S4. Within-group effects on basic target process modulation: VS Transfer**

Points represent Hedges' g for each transfer run (1–2), with error bars indicating 90% confidence intervals. The dashed horizontal line denotes zero effect. Panels display results separately for each group: Group 1 (VS ↓), Group 2 (rIFG ↑), Group 3 (negative VS–rIFG connectivity ↑), Group 4 (A1 ↑), and Group 5 (A1 ↓).

*Table S17 rIFG: Transfer Runs*

| Group | Run | t | df | p | g | 90% CI for g |
|---|---|---|---|---|---|---|
| VS ↓ | Transfer1 | 0.777 | 10 | .455 | 0.216 | [-0.248, 0.680] |
| VS ↓ | Transfer2 | 5.563 | 8 | .001 | 1.675 | [0.858, 2.491] |
| rIFG ↑ | Transfer1 | 1.232 | 11 | .244 | 0.331 | [-0.125, 0.786] |
| rIFG ↑ | Transfer2 | -1.251 | 9 | .242 | -0.362 | [-0.856, 0.132] |
| Neg. VS–rIFG Conn ↑ | Transfer1 | 3.027 | 10 | .013 | 0.843 | [0.298, 1.387] |
| Neg. VS–rIFG Conn ↑ | Transfer2 | 4.856 | 8 | .001 | 1.462 | [0.709, 2.215] |
| A1 ↑ | Transfer1 | 3.004 | 8 | .017 | 0.904 | [0.298, 1.511] |
| A1 ↑ | Transfer2 | 1.740 | 7 | .125 | 0.547 | [-0.017, 1.111] |
| A1 ↓ | Transfer1 | 1.747 | 12 | .106 | 0.454 | [0.002, 0.905] |
| A1 ↓ | Transfer2 | 2.764 | 13 | .016 | 0.695 | [0.228, 1.162] |

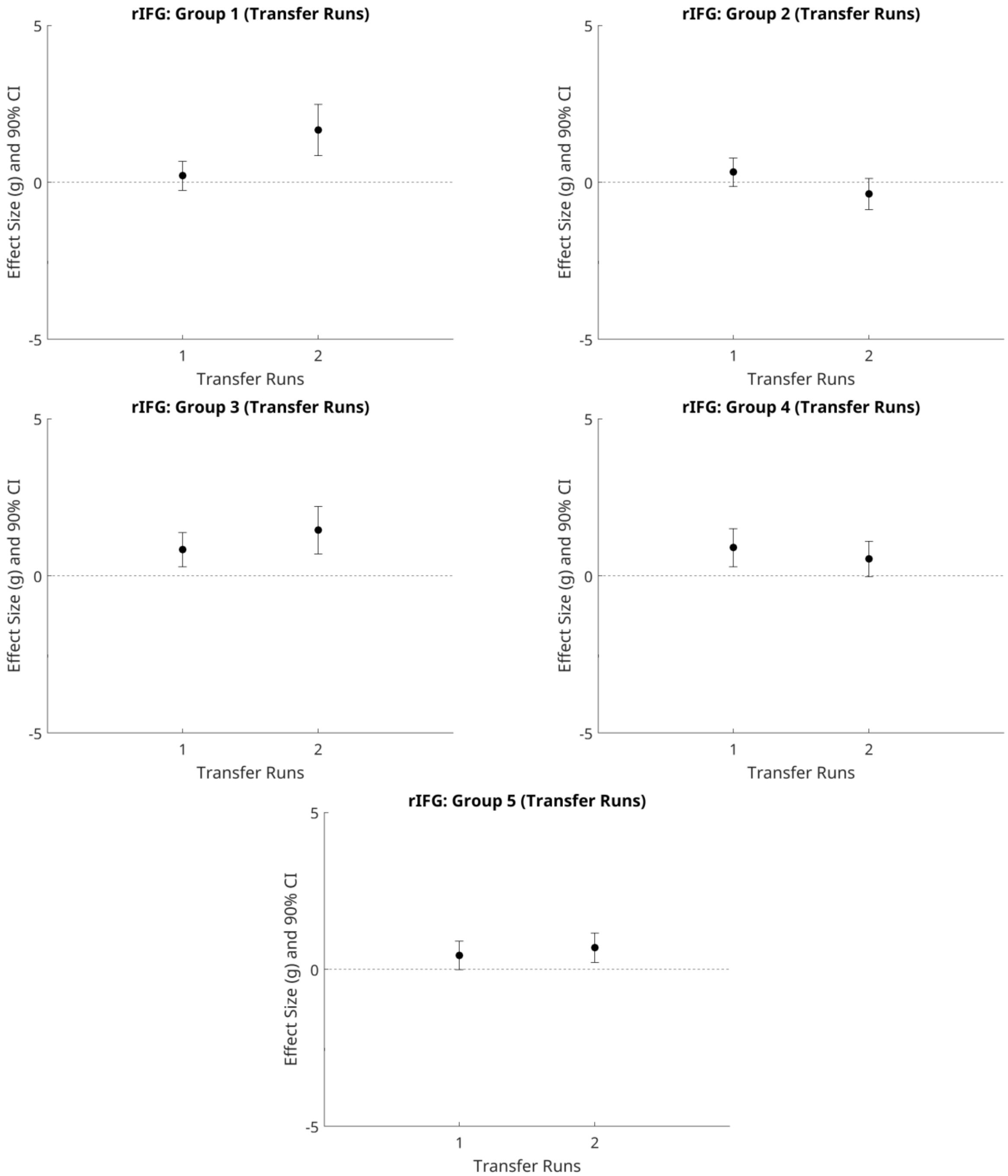


**Figure S5. Within-group effects on basic target process modulation: rIFG Transfer**

Points represent Hedges' g for each transfer run (1–2), with error bars indicating 90% confidence intervals. The dashed horizontal line denotes zero effect. Panels display results separately for each group: Group 1 (VS ↓), Group 2 (rIFG ↑), Group 3 (negative VS–rIFG connectivity ↑), Group 4 (A1 ↑), and Group 5 (A1 ↓).

*Table S18 A1: Transfer Runs*

| Group | Run | t | df | p | g | 90% CI for g |
|---|---|---|---|---|---|---|
| VS ↓ | Transfer1 | -1.849 | 10 | .094 | -0.515 | [-1.007, -0.023] |
| VS ↓ | Transfer2 | 0.112 | 9 | .913 | 0.033 | [-0.443, 0.508] |
| rIFG ↑ | Transfer1 | 0.522 | 12 | .611 | 0.136 | [-0.294, 0.565] |
| rIFG ↑ | Transfer2 | -1.805 | 10 | .101 | -0.502 | [-0.993, -0.012] |
| Neg. VS–rIFG Conn ↑ | Transfer1 | -0.212 | 10 | .836 | -0.059 | [-0.517, 0.399] |
| Neg. VS–rIFG Conn ↑ | Transfer2 | 0.498 | 8 | .632 | 0.150 | [-0.349, 0.649] |
| A1 ↑ | Transfer1 | 1.222 | 8 | .256 | 0.368 | [-0.147, 0.883] |
| A1 ↑ | Transfer2 | -1.013 | 7 | .345 | -0.318 | [-0.852, 0.215] |
| A1 ↓ | Transfer1 | -0.875 | 12 | .399 | -0.227 | [-0.660, 0.206] |
| A1 ↓ | Transfer2 | -1.256 | 14 | .230 | -0.307 | [-0.719, 0.105] |

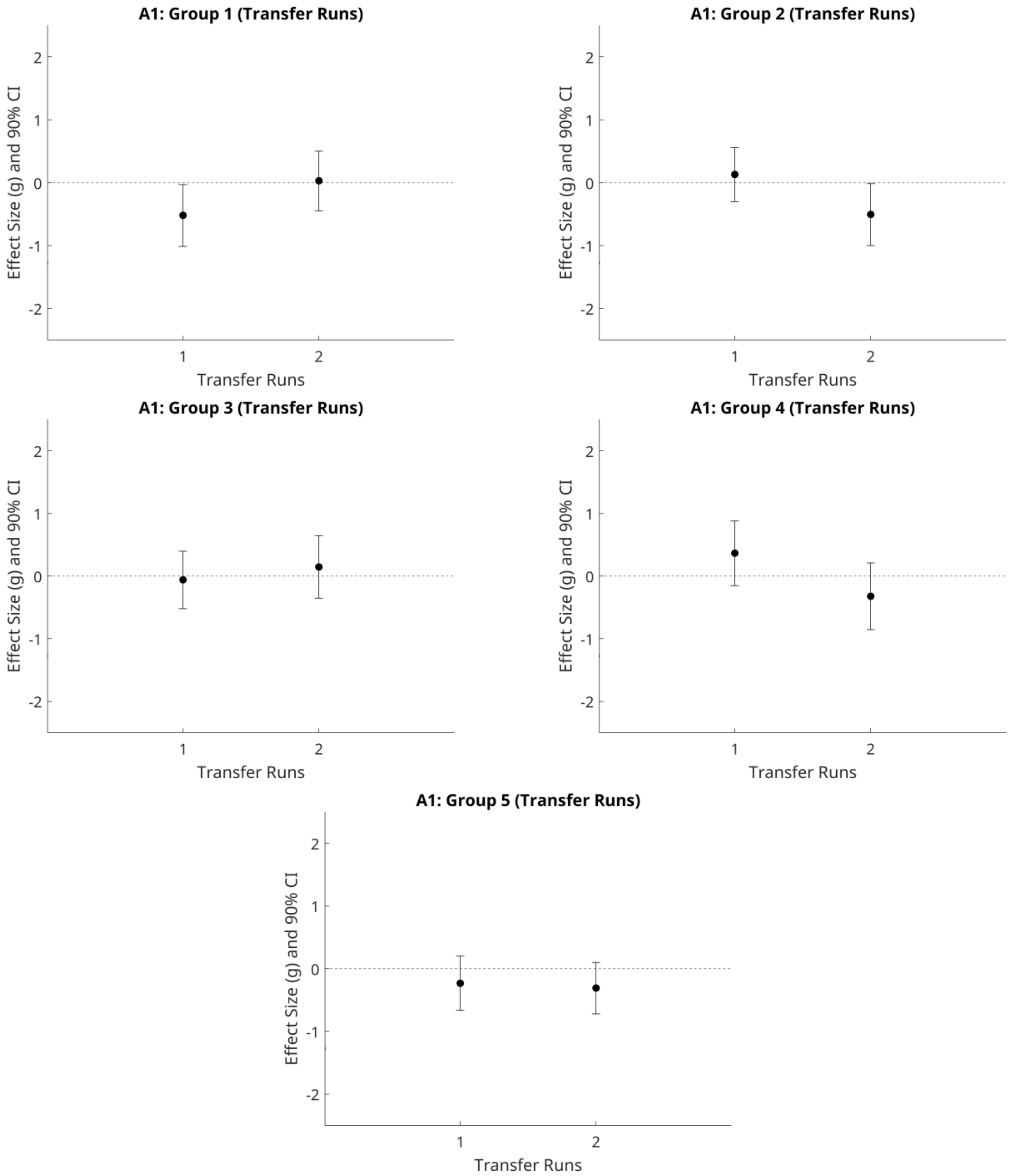


**Figure S6. Within-group effects on basic target process modulation: A1 Transfer**

Points represent Hedges' g for each transfer run (1–2), with error bars indicating 90% confidence intervals. The dashed horizontal line denotes zero effect. Panels display results separately for each group: Group 1 (VS ↓), Group 2 (rIFG ↑), Group 3 (negative VS–rIFG connectivity ↑), Group 4 (A1 ↑), and Group 5 (A1 ↓).

*Table S19 One-sample t-tests examining mean craving change across the three MRI sessions within each intervention group*

| Group | n | M | t(df) | p | 95% CI |
|---|---|---|---|---|---|
| 1 | 10 | 0.54 | 0.66 (9) | .525 | [-1.31, 2.40] |
| 2 | 11 | 0.19 | 0.19 (10) | .852 | [-2.01, 2.39] |
| 3 | 9 | 0.69 | 0.51 (8) | .622 | [-2.43, 3.82] |
| 4 | 8 | -4.74 | -1.78 (7) | .119 | [-11.05, 1.57] |
| 5 | 16 | -1.65 | -1.02 (15) | .326 | [-5.10, 1.81] |

Note: Group 1 (VS ↓), Group 2 (rIFG ↑), Group 3 (negative VS–rIFG connectivity ↑), Group 4 (A1 ↑), and Group 5 (A1). Craving was assessed using four visual analog scale items (0-100). A mean composite score was calculated over scanning days and across items assessing current urge to drink, intention to consume alcohol, expected positive effects of drinking, and expected relief of negative affect or withdrawal symptoms. Positive values indicate an increase in craving from pre- to post-assessment, whereas negative values indicate a decrease. One-sample t-tests were conducted against a test value of zero to determine whether the mean craving change differed significantly from no change.

*Table S20 One-way ANOVA results for post-training craving scores across the five study groups at each MRI session.*

| Outcome Variable | Source | SS | df | MS | F | p |
|---|---|---|---|---|---|---|
| Post MRI1 Craving | Between Groups | 304.82 | 4 | 76.21 | 0.33 | .856 |
| | Within Groups | 13359.07 | 58 | 230.33 | | |
| | Total | 13663.89 | 62 | | | |
| Post MRI2 Craving | Between Groups | 991.97 | 4 | 247.99 | 1.38 | .252 |
| | Within Groups | 9498.90 | 53 | 179.23 | | |
| | Total | 10490.87 | 57 | | | |
| Post MRI3 Craving | Between Groups | 1238.45 | 4 | 309.61 | 1.90 | .125 |
| | Within Groups | 8138.37 | 50 | 162.77 | | |
| | Total | 9376.81 | 54 | | | |

Note: Craving was assessed using four visual analog scale items (0-100). A mean composite score was calculated across items assessing current urge to drink, intention to consume alcohol, expected positive effects of drinking, and expected relief of negative affect or withdrawal symptoms. Higher scores indicated greater craving.

*Table S21 One-way ANOVA results for alcohol consumption outcomes across the five study groups at follow-up assessments.*

| **Outcome Variable** | **Source** | **SS** | **df** | **MS** | **F** | **p** |
|---|---|---|---|---|---|---|
| FU1 Cumulative Alcohol Consumption | Between Groups | 2,731,441.91 | 4 | 682,860.48 | 0.73 | .580 |
| | Within Groups | 32,882,388.87 | 35 | 939,496.83 | | |
| | Total | 35,613,830.78 | 39 | | | |
| FU1 Number of Drinking Days | Between Groups | 67.90 | 4 | 16.97 | 0.86 | .496 |
| | Within Groups | 709.23 | 36 | 19.70 | | |
| | Total | 777.12 | 40 | | | |
| FU2 Cumulative Alcohol Consumption | Between Groups | 1,602,337.60 | 4 | 400,584.40 | 0.25 | .911 |
| | Within Groups | 50,751,924.96 | 31 | 1,637,158.87 | | |
| | Total | 52,354,262.56 | 35 | | | |
| FU2 Number of Drinking Days | Between Groups | 8.98 | 4 | 2.24 | 0.05 | .996 |
| | Within Groups | 1,503.33 | 31 | 48.49 | | |
| | Total | 1,512.31 | 35 | | | |
| FU3 Cumulative Alcohol Consumption | Between Groups | 33,400,590.65 | 4 | 8,350,147.66 | 1.13 | .367 |
| | Within Groups | 185,528,894.71 | 25 | 7,421,155.79 | | |
| | Total | 218,929,485.37 | 29 | | | |
| FU3 Number of Drinking Days | Between Groups | 125.38 | 4 | 31.35 | 0.37 | .826 |
| | Within Groups | 2,100.79 | 25 | 84.03 | | |
| | Total | 2,226.17 | 29 | | | |

Note. One-way analyses of variance examined follow-up alcohol consumption outcomes across the five neurofeedback/control groups at three follow-up assessments (FU1, FU2, FU3). Outcomes included cumulative alcohol consumption and number of drinking days. No significant between-group differences were observed for any follow-up outcome. SS = sum of squares; MS = mean square; FU = follow-up.
